# Towards One Health surveillance of antibiotic resistance, use and residues of antibiotics in France: characterisation and mapping of existing programmes in humans, animals, food and the environment

**DOI:** 10.1101/2022.11.14.22281639

**Authors:** L. Collineau, C. Bourély, L. Rousset, A. Berger-Carbonne, MC. Ploy, C. Pulcini, M. Colomb-Cotinat

## Abstract

International organizations are calling for One Health approaches to tackle antibiotic resistance (ABR). In France, the diversity of surveillance programmes makes it difficult to get an overview of the current surveillance system and its level of integration. This study aimed to map and characterise all French surveillance programmes for ABR, antibiotic use (ABU) and antibiotic residues in humans, animals, food and the environment, to identify integration points, gaps and overlaps.

A literature review and interviews with 36 programme coordinators were conducted to identify and characterise programmes using a standardized grid (28 variables). Forty-eight programmes were included. They targeted the human (n=35), animal (n=12), food (n=3) and/or the environment (n=1); 35 programmes focused on ABR, 14 on ABU and two on antibiotic residues. Two programmes were cross-sectoral. Among the 35 ABR programmes, 23 collected bacterial isolates. Bacteria most targeted were *Escherichia coli* (n=17 programmes), *Klebsiella pneumoniae* (n=13), and *Staphylococcus aureus* (n=12). ESBL-producing *E. coli* was monitored by the majority of ABR programmes in humans, animals and food, and is a good candidate for integrated data analysis. ABU indicators were highly variable. Areas poorly covered were the environmental sector, overseas territories, ABR colonisation in humans and ABU in companion animals.

The French surveillance system appears rich and extensive, but with gaps and only few integration points. We believe this mapping will be of high interest to policy makers and surveillance stakeholders, and that our methodology may inspire other countries willing to progress towards One Health surveillance of ABR.

## Introduction

Antibiotic resistance (ABR) is a threat to modern health care and is recognized as one of the major public health problems [1, 2]. Since antibiotic-resistant microorganisms exist in all ecosystems, with transmission of bacteria and resistance genes, prevention of ABR is a complex issue, which requires development of integrated policies at the human-animal-environment interface. Therefore, the WHO Global Action Plan on Antimicrobial resistance [3], jointly adopted by the World Organisation for Animal Health and the Food and Agriculture Organization of the United Nations, underscored the need for surveillance using a One Health approach. The 2017 EU One Health Antimicrobial Resistance Action Plan [4] also argued for a more integrated surveillance system of antibiotic resistance, including closely related topics, namely antibiotic use (ABU) and antibiotic residues in ecosystems. In Europe, most countries have already set up mandatory or voluntary surveillance programmes in humans [5], companion and food-producing animals and food [6]. The joint inter-agency report on integrated analysis of antimicrobial agents consumption and occurrence of antimicrobial resistance in bacteria from humans and food-producing animals (JIACRA) contributes to better understand ABR and provides valuable insights for policy-makers across the EU [7].

In France, numerous surveillance programmes are currently in place, covering ABR and ABU in both humans and animals, as well as antibiotic residues. Under the impulsion of the 2016 interministerial roadmap for controlling antimicrobial resistance [8], there is a will to rationalize surveillance data across sectors and to promote cross-sectoral collaborations, in addition to sectorial national action plans [9-11]. However, the large number and diversity of surveillance programmes make it difficult to have an exhaustive picture of the overall surveillance system. A comprehensive mapping of existing programmes is currently lacking. This essential first step will facilitate the ongoing evaluation of the collaborations between surveillance programmes and identification of key levers to improve cross-sectoral collaboration.

Hence, the aim of this study was to identify, map and characterise all French surveillance programmes for ABR, ABU and antibiotic residues existing in humans, animals, food and the environment, and to identify integration points, gaps and overlaps.

## Methods

### Inclusion and exclusion criteria

As the French surveillance initiatives were highly variable in terms of geographic scope, objectives and sustainability, we retained for analysis only those initiatives corresponding to the following definition of a surveillance programme: “a structured group of actors and/or institutions in charge of collecting, centralizing, analysing and communicating quantitative data on a regular and long-term basis” [12]. Both local/regional and national surveillance programmes were included. Exclusion criteria were unrepeated research studies, inactive programmes at the time of the literature review, clinical research programmes, as well as programmes assessing appropriateness of antibiotic use. The focus was on antibiotics only, excluding other antimicrobials.

### Identification of surveillance programmes

A literature review was conducted in January-February 2021 in both the scientific and grey literatures (in English and French languages), to identify all potential French programmes for surveillance of ABR, ABU and antibiotic residues in human, animal, food and the environment. Primary literature sources were official websites of ministries, public health agencies, and other public and private institutions involved in ABR-related surveillance. To screen the scientific literature, the following search string was used in PubMed^®^, including articles published since 2005 only: (antimicrobial*[Title/Abstract] OR antibiotic*)[Title/Abstract] AND (surveillance[Title/Abstract] OR monitoring)[Title/Abstract] AND France[Title/Abstract]. After listing all potential programmes identified, the coordinator of each programme was contacted by email to check if the programme matched the inclusion criteria. Lastly, the list of identified programmes was submitted to a group of 20 French experts with long-term expertise in surveillance or policy-making related to ABR, ABU and antibiotic residues surveillance in the human, animal, food and environmental sectors, in order to identify any potential missing programme and validate the final list.

### Characterisation of surveillance programmes and mapping

Surveillance programmes were characterised using a standardised grid adapted from the ECoSur matrix developed by Bordier et al. [13]. The grid included 28 variables of interest covering aspects related to organisation (e.g. regulatory status, ownership, steering and coordination activities), methods and operations (e.g. target population, coverage, sampling strategy, data collection and analysis, indicators used, dissemination of the results). Contribution to supra-national surveillance programmes was also recorded. A detailed list of collected variables is provided in Supplementary material (Table S1).

For each included surveillance programme, the descriptive grid was pre-completed using information available from the literature collected by the research team, made of two scientists from the human sector and two from the animal sector. Then, interviews with the programmes’ coordinators were conducted by the research team to complete and validate the grid. Interviews were performed between February and June 2021, using online videoconferencing because of Covid-19 related restrictions. Lastly, based on collected data, a visual representation was produced to display the mapping of the surveillance system and make it easier to identify integration points across sectors, as well as overlaps and gaps.

## Results

### Selection process and data collection

Out of the 79 surveillance initiatives initially identified, 48 matched our inclusion criteria and were included in further analysis (see Table S2 in supplementary material). A total of 36 interviews with programme coordinators was conducted to collect information about 40 programmes. For the remaining eight programmes, information was validated via email exchange.

### Sectors, populations and targets

Out of the 48 included programmes, 35 targeted the human, 12 the animal, three the food and one the environmental sectors (Table 1). Two programmes were cross-sectoral, and covered both the human and animal or food sectors (Figure 1). In the human sector, seven national programmes belonged to the French network for prevention of healthcare-associated infections and ABR (RéPias), launched in 2018 and led by Santé publique France, the French public health agency. The Répias is a key support to the national strategy for preventing infections and ABR in the human sector; it produces surveillance data on healthcare associated infections, ABR and antibiotic consumption, and supports infection prevention and control tools and public health communication media [14]. In addition, the French National Observatory for Epidemiology of Bacterial Resistance to Antimicrobials (ONERBA), existing since 1997, grouped together eight voluntary programmes, of which four were bacterial species-specific, and two had regional coverage [15]. Last, among the 35 programmes targeting humans, 14 were National Reference Centres (NRCs) coordinated by Santé publique France.

**Table 1.** Distribution of surveillance programmes according to sector, population and target (n=48 programmes)*, France, 2021.

| Sector<br>(n=number of<br>programmes) | Population (n=number<br>of programmes) | Number of programmes covering the target of<br>interest** (IDs of corresponding programmes) |  |  |
| --- | --- | --- | --- | --- |
|  |  | ABR (n=35) | ABU (n=14) | Residues<br>(n=2) |
| Human (n=35) | Healthcare facilities<br>(n=30) | 29 (1-17, 19, 21,<br>23-24, 28, 32-36,<br>44, 46) | 3 (18, 21, 44) | not<br>applicable |
|  | Community (n=23) | 19 (1, 3-16, 19, 24,<br>35, 45) | 4 (18, 37, 39,<br>48) | not<br>applicable |
|  | Long-term care facilities<br>(n=20) | 18 (3-16, 20, 24,<br>35, 45) | 3 (18, 20, 37) | not<br>applicable |
| Animal (n=12) | Diseased food-producing animals<br>(n=10) | 3 (25, 30-31) | 7 (22, 27, 38,<br>40-43) | not<br>applicable |
|  | Diseased companion<br>animals<br>(n=2) | 1 (30) | 1 (43) | not<br>applicable |
|  | Healthy food-producing<br>animals (n=2) | 2 (19, 26) | none | not<br>applicable |
| Food (n=3) | Food of animal and non-<br>animal origin (n=1) | 1 (11) | none | none |
|  | Food of animal origin (n=2) | 1 (26) | none | 1 (29) |
| Environnement (n=1) | Surface and ground water (n=1) | none | none | 1 (47) |

*\* A programme may target more than one sector, population or target*
*\*\* To see the correspondence between identifiers (IDs) and names of the programme, see Supplementary material*

**Figure 1.**
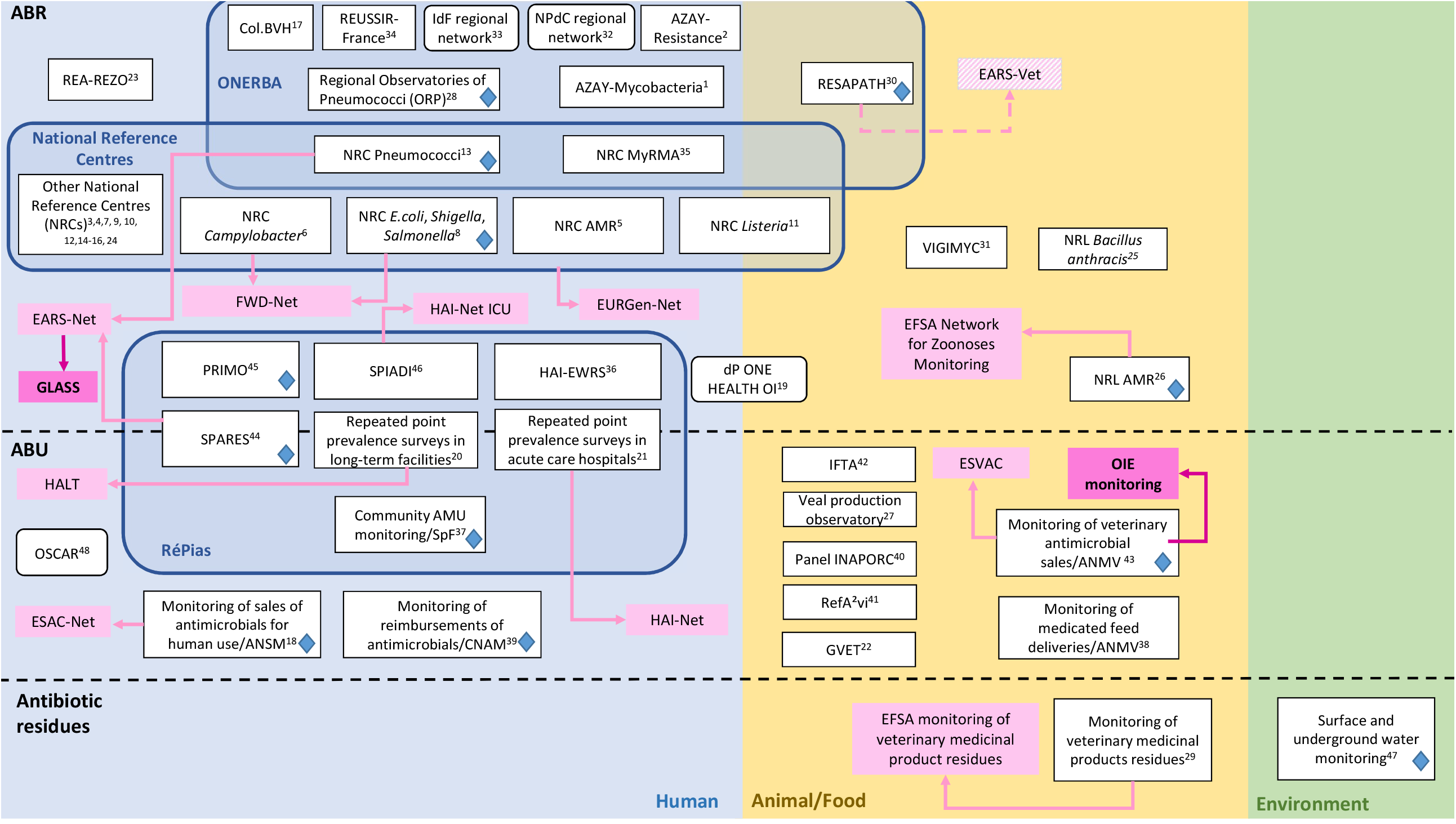
Mapping of the existing surveillance programmes for antibiotic resistance (ABR), antibiotic use (ABU) and antibiotic residues in humans, animals/food and the environment in France in 2021 White boxes: French surveillance programmes (straights corners = national; rounded corners = regional). Light pink boxes: European surveillance programmes (with EARS-Vet under construction). Dark pink boxes: international surveillance programmes. Blue diamonds: programmes contributing to the annual joint One Health Antibiotic Resistance brochure coordinated by Santé publique France at the occasion of the annual World Antimicrobial Awareness Week (12 programmes involved). Superscripts numbers correspond to programme IDs detailed in supplementary material.

Among the 48 programmes, the majority (n=35) focused on ABR (Table 1). Three programmes in the human sector monitored both ABR and ABU. All 31 human ABR-related programmes collected data from clinical samples; two programmes also collected data from screening samples (colonisation). In the animal sector, the majority of programmes focused on diseased or healthy food-producing animals (n=12) and targeted multiple animal species (n=7), horses (n=3), pigs (n=2), poultry (n=1), veal calves (n=1) or rabbits (n=1). Two programmes targeted diseased companion animals, including dogs (n=2) and cats (n=2). The environmental programme targeted surface and ground water.

### Regulatory status, funding and durability

The French surveillance system relied mainly on public funds, as 34 out of 48 programmes were publicly funded. Nine programmes were based on mixed public-private funding, and five were privately funded (Table S1). Surveillance programmes were in majority regulated (coordinated by authorities, but implemented by other actors) or official (coordinated and implemented by authorities) (n=29), although 19 programmes were independently run by voluntary actors. The majority of programmes (n=27) were established before 2010, and 15 were built within the last 10 years, of which nine in the last five years. For six programmes, the creation date could not be retrieved.

### Timeliness, geographic coverage and granularity

Most programmes (n=24) collected data throughout the year without interruption, although some programmes collected data annually (n=14), infra-annually (n=6) or pluri-annually (e.g. every three years, n=3). Dissemination of the results occurred on an annual (n=43), pluri-annual (n=3) or infra-annual (n=2) basis. Sixteen out of 48 programmes also disseminated their results via open-access dashboards. Most programmes had national coverage (n=43) and among them, 31 included at least one overseas territory. Among the 43 nationwide programmes, all displayed their results at a national level, but 10 programmes with a higher granularity also displayed their results at a sub-national level. In addition, five local programmes displayed their results at a single regional level only.

### Surveillance design and data collected

The majority (41/48) of surveillance programmes relied on passive surveillance. Among the 35 ABR surveillance programmes, 23 had access to bacterial isolates and were able to perform molecular characterisation (e.g. PCR or whole-genome sequencing) on all or part of the collected isolates in addition to conventional antimicrobial susceptibility testing (Table 2). ABU surveillance was primarily based on administration (n=6), deliveries/dispensing data (n=5), reimbursements (n=3) or sales data (n=2).

**Table 2.** Distribution of surveillance programmes according to data collected (n=48 programmes)*, France, 2021.

| Target | Number of programmes (n=) | Data collected | Number of programmes (n=) | Corresponding IDs** |
| --- | --- | --- | --- | --- |
| ABR | 35 | Resistance data*** only | 12 | 2, 17, 20-21, 23, 32-34, 36, 44-46 |
|  |  | Resistance data and bacterial isolates | 23 | 1, 3-16, 19, 24-26, 28, 30, 31, 35 |
| ABU | 14 | Administration | 6 | 20-22, 27, 41-42 |
|  |  | Deliveries or dispensing | 5 | 27, 38, 40-41, 44 |
|  |  | Reimbursements | 3 | 37, 39, 48 |
|  |  | Sales | 2 | 18, 43 |
|  |  | Prescriptions | 1 | 41 |
| Residues |  | Residues data | 2 | 29, 47 |

\* A programme may collect several types of data
\*\* To see the correspondence between identifiers (IDs) and names of the programme, see Supplementary material
\*\*\* Resistance data include either quantitative (minimum inhibitory concentrations or disk diffusion diameters) or qualitative (SIR: susceptible-intermediate-resistant profiles) data from antimicrobial susceptibility testing, without further access to bacterial isolates

### Targeted bacteria

Among the 35 programmes monitoring ABR in humans or animals, 19 covered multiple bacterial species simultaneously, while 16 targeted a single bacterial species (Table S1), typically programmes run by NRCs (n=14). Bacterial species of primary interest were *Escherichia coli* (n=17 programmes), *Klebsiella pneumoniae* (n=13), *Staphylococcus aureus* (n=12). More specifically, extended-spectrum beta-lactamases (ESBLs)-producing *E. coli* were monitored by 17 programmes and methicillin-resistant *Staphylococcus aureus* (MRSA) by 12 programmes (Table 3).

**Table 3.**
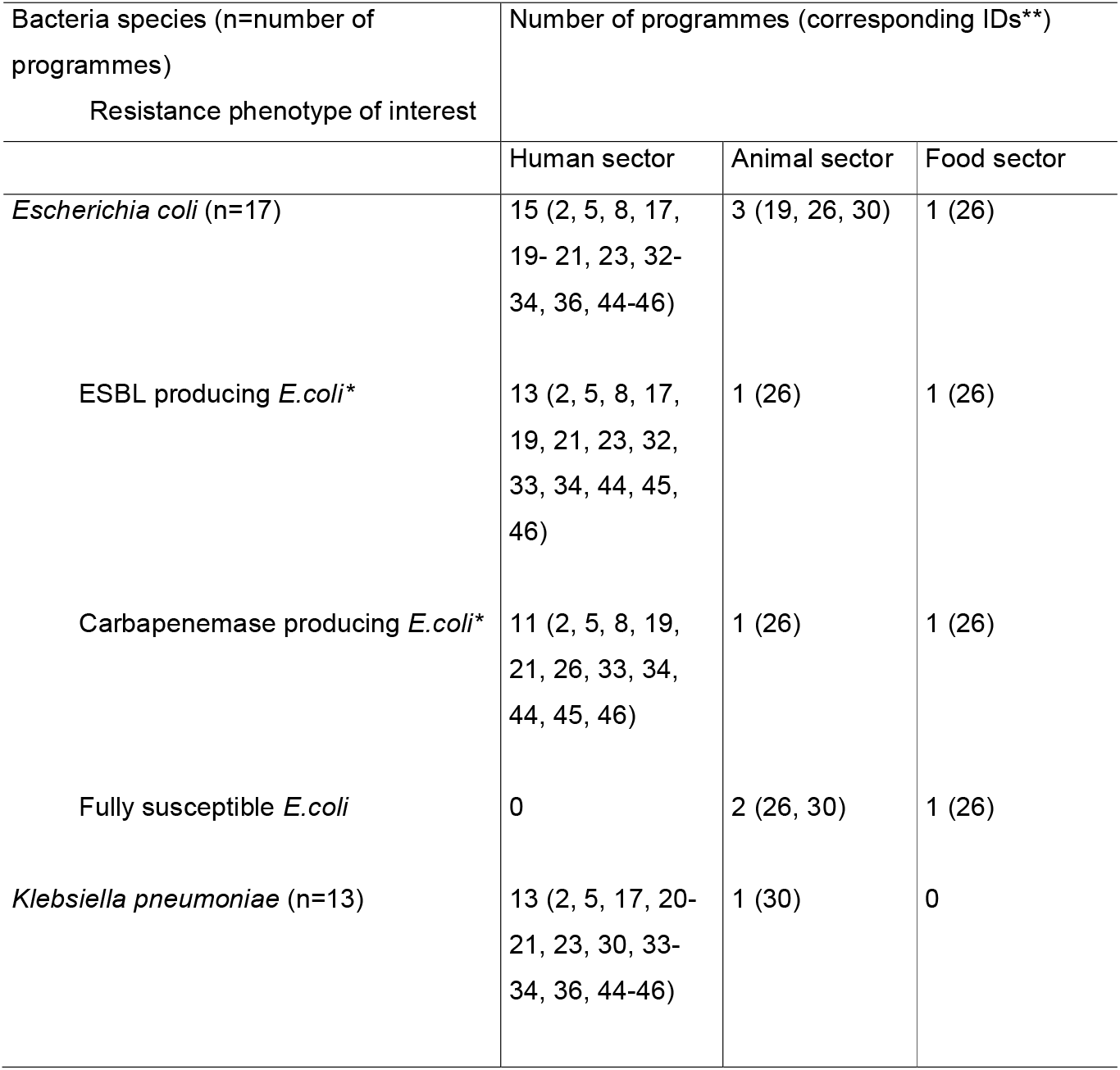
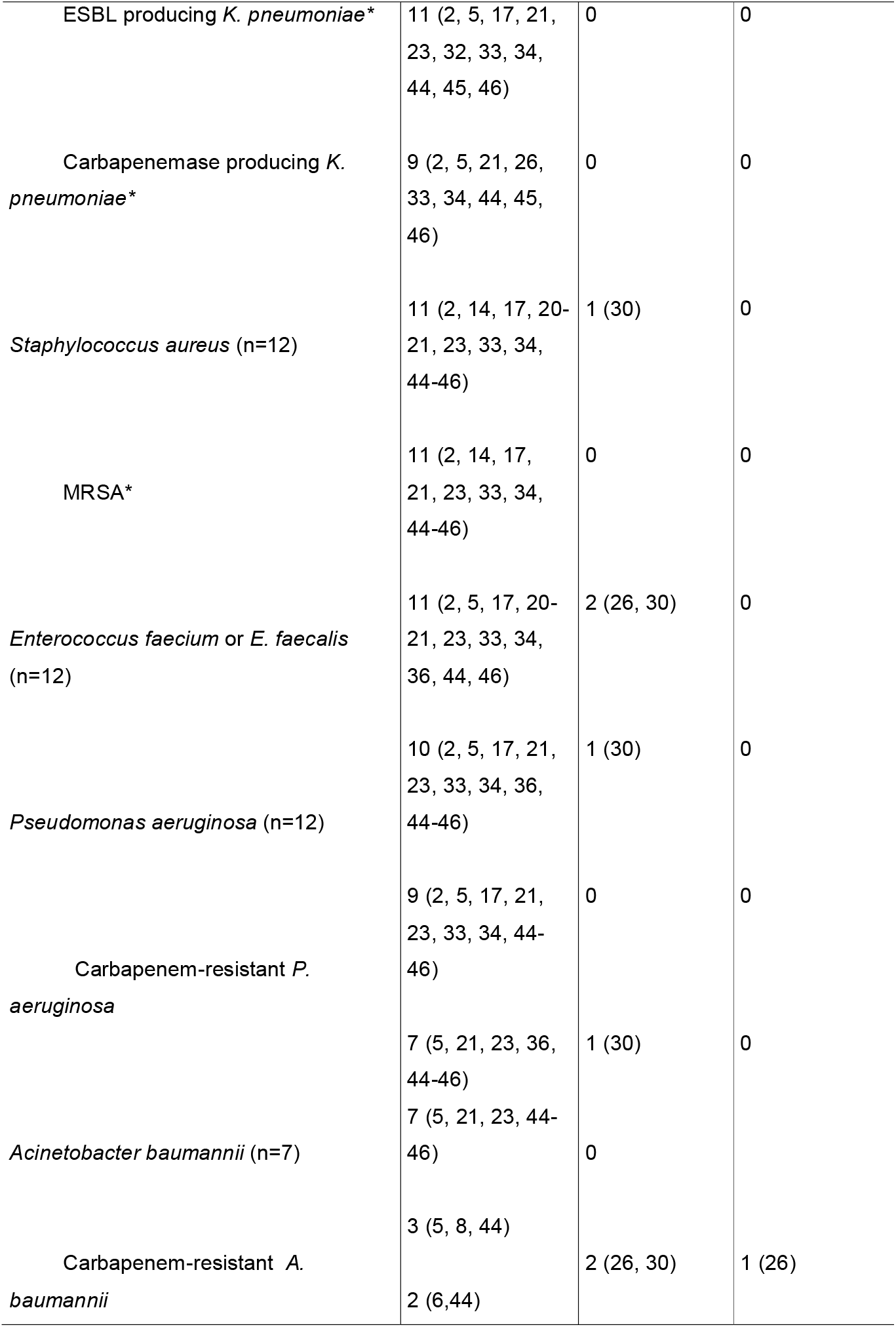
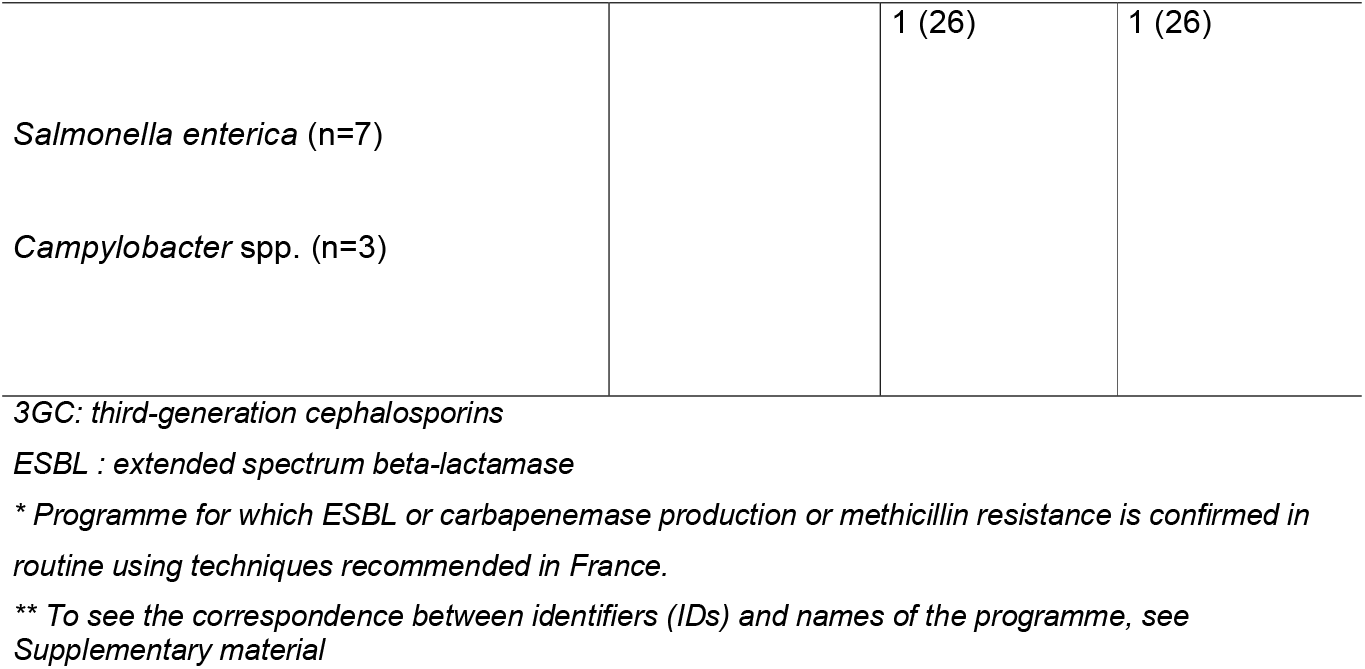
Bacteria species and resistance phenotypes most commonly monitored in the human, animal and food sector, France, 2021.

### Indicators of ABR, ABU and antibiotic residues

The large majority of ABR-related programmes monitored a proportion of resistant isolates (33/35), although a few programmes worked with different indicators, including the incidence rate (n=4) or number of cases (n=2) of infections with ABR bacteria, as well as the prevalence of samples harbouring at least one ABR isolate (n=1). Different standards were used to determine resistance profiles: all 31 human-related programmes were using clinical breakpoints defined by the European Committee on Antimicrobial Susceptibility Testing (EUCAST), animal-related programmes were using epidemiological cut-off values (ECOFFs) from either EUCAST (n=2) or the veterinary section of the Antibiogram Committee of the French Society of Microbiology (CASFM) (n=1), or clinical breakpoints from the Clinical and Laboratory Standards Institute (CLSI) (n=1) or simply providing minimum inhibitory concentration distributions in the absence of available interpretation criteria (n=1, *Mycoplasma* spp. in ruminant animals). The indicators for ABU surveillance were highly variable both within and between the human and animal sectors (Table 4) and depending on the targeted population. Detailed description of ABU indicators calculation has been provided elsewhere [16].

**Table 4.**
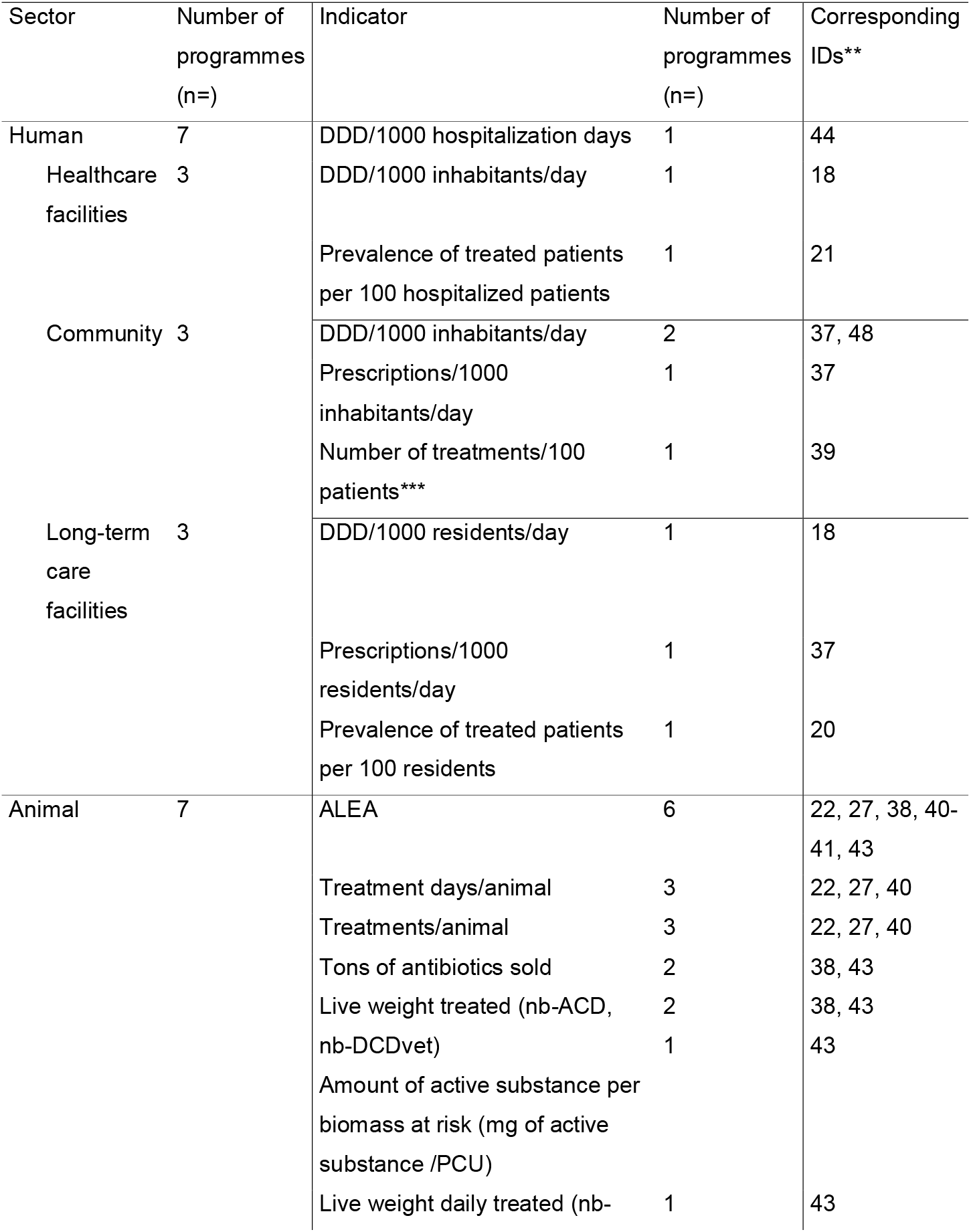
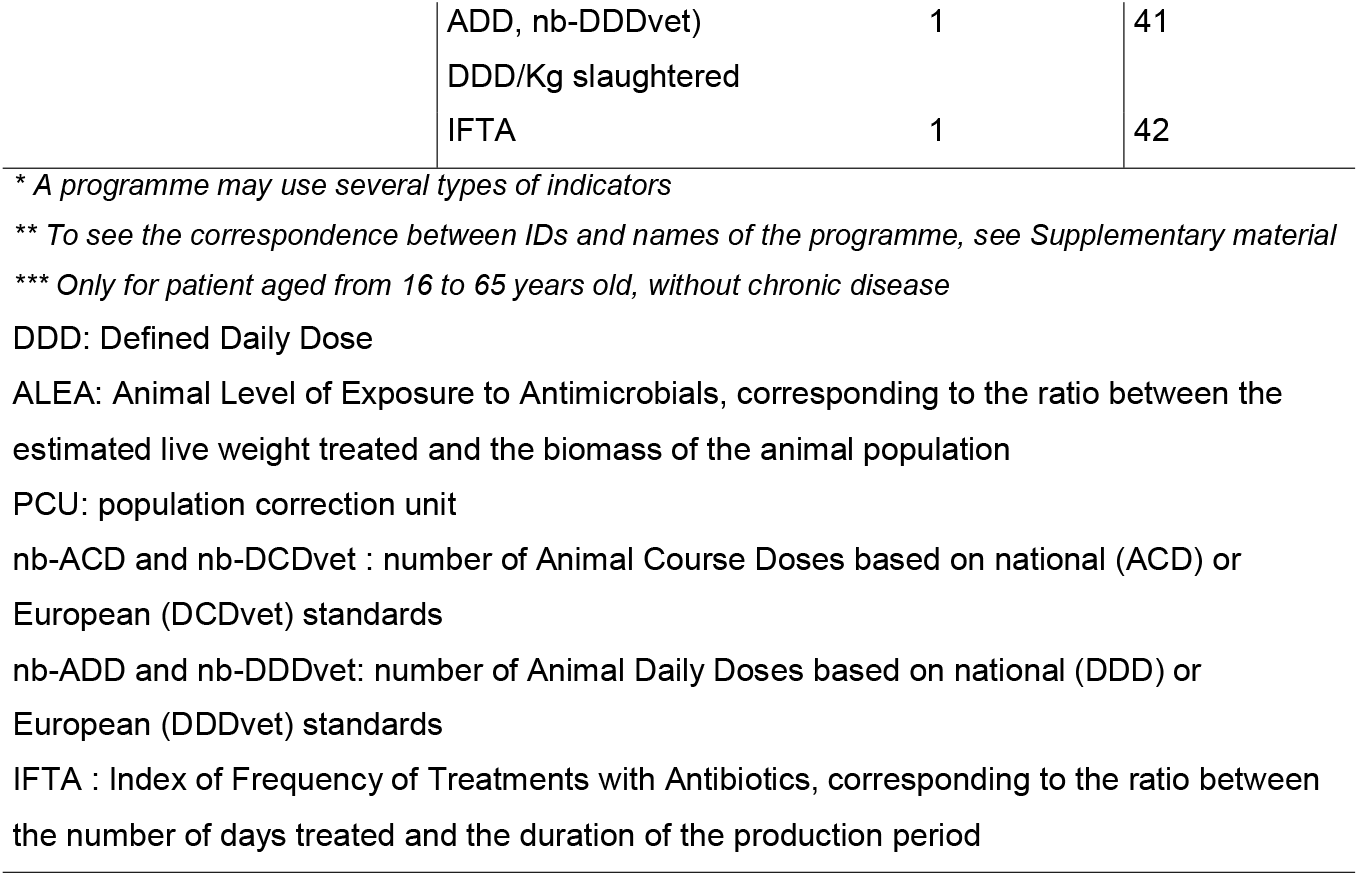
Distribution of surveillance programmes according to ABU indicators (n=14 programmes)*, France, 2021.

Regarding antibiotic residues surveillance, the indicator used in one food programme was the proportion of samples beyond the maximum residue level (MRL) for sulphonamides and quinolones [17], while the indicator used in the environment was the proportion of samples beyond the Predicted No Effect Concentration (PNEC) for macrolides, fluoroquinolones and sulphonamides-diaminopyrimidines [18].

### Contribution to supranational surveillance programmes

In addition to supporting national initiatives against ABR, the French surveillance programmes contributed to ten European and two international established programmes for surveillance of ABR, ABU, or antibiotic residues (Figure 1), and to one programme under construction for ABR surveillance in diseased animals (EARS-Vet) [19].

## Discussion

The present study provided the first comprehensive overview of the French surveillance system for ABR, ABU, and antibiotic residues, including a mapping (Figure 1) and detailed characterization of the 48 surveillance programmes existing in 2021 in humans, animals, food, and the environment, as well as the identification of major gaps and overlaps (Box 1). For comparison, 11 programmes for ABR/ABU surveillance were identified in the UK [20] ; 29 in Canada including six national, 22 provincial and one territorial programme [21]. The large number of French programmes was partly related to several characteristics: i) epidemiological and molecular ABR data were collected through separate surveillance programmes, ii) NRCs in humans and National Reference Laboratories (NRLs) in animals were split by bacterial species, and iii) ABU surveillance programmes were built by separate animal species in the animal sector.

Despite their large number, the French programmes complemented each other by targeting different populations and providing evidence to support and evaluate national actions [9-11]. Moreover, the majority of programmes produced surveillance reports at least annually, which appeared sufficient to support operational surveillance of the ABR epidemiological situation and guide prevention and control strategies. Several programmes also contributed to supra-national programmes, hence facilitating the surveillance system to respond to EU and international requirements.

However, the French surveillance system appeared fragmented, as the large majority of surveillance programmes were addressing a single sector, and focused on either ABR or ABU. Only three programmes, all in the human sector, targeted both ABR and ABU. Moreover, only two programmes covered both human and animal sectors, with one programme being local.

Similar to France, the UK surveillance system appeared fragmented with limited integration between surveillance programmes. Conversely, the Canadian ABR Surveillance System appeared at an advanced stage of integration, although surveillance coverage was incomplete and highly variable between provinces/territories.

In France, two subsystems partly counterbalanced the apparent lack of integration, by facilitating collaborations between programmes. The RéPias contributed to integrate ABU and ABR in the human sector, while ONERBA facilitated ABR integration between the human and animal sectors. In addition, a joint One Health Antibiotic Resistance brochure [22], led by Santé publique France and gathering 12 programmes from the three sectors, is published each year during the World Antimicrobial Awareness Week. It appeared as an integrative One Health effort across sectors and targets, although limited to joint external communication of the results produced independently by each programme. An additional working group facilitating integrated analyses across sectors, inspired from JIACRA but based on French-specific data, would nicely complement this activity.

Third-generation cephalosporins resistant *E. coli*, and especially ESBL-producing *E. coli*, was monitored by the majority of ABR surveillance programmes in human, animal and food sectors, and appeared as a good candidate for integrated data analysis, hence complementing sectoral monitoring, as already suggested by ongoing One Health initiatives on antibiotic resistance, such as the WHO Tricycle protocol [23]. Conversely, ABU indicators showed large variations within and between sectors, which could hinder data integration efforts and interoperability. ABR indicators were more harmonized, but the interpretation criteria and antibiotic susceptibility testing standards differed.

A few overlaps were identified in the French surveillance system, mainly in the human sector, where five programmes targeted ABR in healthcare facilities. These overlaps are due to older programmes at local or national levels that persisted when the programmes of the RéPias were initiated in 2018. There is a need to clarify objectives of these overlapping programmes, in order to improve the visibility and efficiency of the system.

Conversely, we pointed out several gaps in the French surveillance system. First, the environmental sector was largely uncovered: we identified only one programme that complied with our definition of a surveillance programme. Other initiatives existed but were not sustainable at this stage. Structured national surveillance of antibiotic residues was limited to surface water and animal-derived food. No residues surveillance programme was identified in other important areas such as farm environments or wastewater treatment plants, although various research studies explored this issue (not presented here). This was not surprising as worldwide efforts towards environmental surveillance of ABR and antibiotic residues have recently started [24]. Still, the inclusion of the environmental sector in One Health approaches has been growing lately, as shown by the recent integration of the United Nations Environment Program (UNEP) into the One Health antimicrobial resistance activities of the Quadripartite Alliance [25]. Of note, the EU watch list for water surveillance targeted only a limited number of antibiotic classes [18]. There is a need to enlarge and strengthen the structuration of ABR and antibiotic residues surveillance in the environment, and to harmonize surveillance indicators being used in this sector, as recommended in the French national action plan for the environment and health [11].

Second, the coverage of the surveillance system could be further improved both in the human and animal sectors. Overseas territories were poorly represented in programmes with national geographic coverage. In the human sector, surveillance covered the three main populations of interest (healthcare facilities, long-term care facilities and the community). However, most human programmes focused on clinical samples reflecting suspicions of infections, with a lack of data on colonisation by antibiotic-resistant bacteria. This may underestimate spread of emerging resistance, e.g. carbapenemase-producing Enterobacterales, for which infection rates remain low in France, but dissemination is increasing [26].

In the animal sector, both healthy and diseased animals were covered by national ABR surveillance programmes, a situation still uncommon in Europe [27]. Additionally, several farm-level ABU surveillance programmes dedicated to selected livestock species complemented the overall surveillance of sales data. Yet, an important gap was a dedicated ABU-surveillance programme in companion animals. However, the upcoming implementation of the EU Regulation 2019/6 on veterinary medicinal products should address this gap within a few years (by 2027 for horses and 2030 for dogs/cats [28]).To meet with this new regulation, a new data collection system will be implemented in France from 2023 onwards; this may challenge the relevance and sustainability of the existing farm-level ABU surveillance programmes. Another gap in the animal sector is the lack of ABR surveillance in non-captive wild animals and aquaculture, although the RESAPATH programme already covers parts of antibiotic susceptibility testing performed in fish production (ref). Additionally, antibiotic susceptibility testing in diseased animals was limited to antimicrobials authorized in veterinary medicine, which limits the assessment of the zoonotic exposure to ABR of human health relevance (e.g. resistance to carbapenems). The EARS-Vet initiative, launched during the EU-JAMRAI European Joint Action (eu-jamrai.eu), which intends to develop a European programme for surveillance of ABR in clinical pathogens of animals, recently proposed a panel of antibiotics of primary interest to both animal and human health [29].

A major strength of this study was the comprehensive approach we used, addressing ABR from a broad perspective including ABR, ABU and antibiotic residues in humans, animals and the environment, since these are closely connected. To our knowledge, no such overview is available elsewhere in the literature. By direct exchange with the coordinators of each programme, we are confident our data are accurate and validated. We believe this mapping will be of interest to policy makers, as well as surveillance stakeholders, not only in France but also in other countries, since our methodology can easily be transferred to other countries and situations.

Nonetheless, this study also had some limitations. While we collected detailed data on ABR/ABU indicators together with other information being generated by each programme, getting access to the actual programme databases, e.g. to look at data formats, or thesaurus, was beyond the scope of this study. Hence, we were unable to evaluate the inter-operability of existing data. Additionally, our mapping only provided a snapshot of the surveillance system in 2021 and did not capture changes over time. Yet, the French ABR-related surveillance system appeared as an ever-evolving system, with several programmes and sub-systems that were launched and others discontinued in the recent years. As an example, two large national meta-networks [30] funded through the French Priority Research Programme on ABR were launched in November 2021: (i) the meta-network PROMISE aims to build a One-Health community of actors on ABR, to develop a joint data warehouse for ABR surveillance and to set up a national network for environmental surveillance of ABR, and (ii) the meta-network ABRomics-PF aims to build a platform for ABR multi-omics One Health data sharing. Those two meta-networks appear as excellent opportunities to further facilitate integration of surveillance programmes, and address some of the gaps identified in this study.

Our study was the first step for the assessment of the ‘One Health-ness’ of the French surveillance system. An in-depth investigation of existing collaborations between surveillance programmes as well as the main drivers for collaboration is still under progress. It will complement our practical recommendations to improve One Health surveillance of ABR in France. We believe that the whole approach of identification and characterisation of surveillance programmes, mapping of the surveillance system to identify gaps and overlaps, and ultimately the evaluation of collaborations between programmes, is an added value to the ABR surveillance landscape and will inspire other countries willing to progress step by step towards One Health surveillance of ABR.

### Box 1

**list of the major gaps and overlap identified in the coverage of the surveillance system for antibiotic resistance, antibiotic use and antibiotic residues, France, 2021**.

#### Gaps

- Lack of structured national surveillance programmes in the environmental sector.
- Antibiotic residues only routinely monitored in surface water and animal-derived food. Overseas territories poorly represented.
- ABR surveillance in the human sector mostly targeting clinical samples, and rarely
- screening samples.
- Lack of a dedicated ABU-surveillance programme in companion animals.
- Lack of ABR surveillance in non-captive wild animals and aquaculture.
- Lack of ABR testing in diseased animals to antibiotics of primary interest in human
- health, since routine testing is limited to antimicrobials authorized in veterinary medicine.

#### Overlap

- Five programmes targeted ABR data collection in healthcare facilities in the human sector.

## Conclusion

This first mapping and characterization of the French surveillance system for ABR, ABU and antibiotic residues showed a rich and varied yet complex and fragmented surveillance system, involving multiple programmes. Overall, these programmes provide good coverage of key target populations in the human and animal sectors; however, some gaps were identified, notably in the environmental sector, which is largely uncovered. This study is an important step for future evaluation of the One health-ness of the French ABR surveillance system.

## Data Availability

All data produced in the present work are contained in the manuscript

## Notes

### Competing Interest Statement

The authors have declared no competing interest.

### Funding Statement

This study was funded by the French Ministry of Agriculture and Food
through the EcoAntibio2 programme (research grant 2019-124). The funder contributed to
data collection but had no role in data analysis, interpretation or communication of the
results.

